# Interprofessional Readiness and Team Climate in Psychiatric Care: Profession-Specific Differences in a Japanese Psychiatric Hospital

**DOI:** 10.1101/2025.11.20.25340704

**Authors:** Yasuhisa Nakamura, Mayumi Yoshikawa, Koshi Terashita, Miki Fukushima, Chizuru Tsubonouchi

## Abstract

Interprofessional collaboration is critical for integrated psychiatric care, yet few studies have profiled clinicians’ readiness for interprofessional learning and team climate across professions while adjusting for demographic and experiential factors. We conducted a single-site cross-sectional survey of physicians, nursing staff, and allied health professionals in a specialist psychiatric hospital in Japan. Primary outcomes were the Readiness for Interprofessional Learning Scale–Professional version (RIPLS-Pro) total score and the Participative Safety subscale of the Team Climate Inventory. We compared professions and clinical settings using regression models adjusted for age and years of practice and analyzed free-text responses using directed content analysis. Profession was associated with both RIPLS-Pro and Participative Safety scores: allied health staff scored the highest, followed by physicians and nursing staff. Clinical setting and the profession-by-setting interaction did not explain additional variance. Age correlated positively with RIPLS-Pro scores. Qualitative data highlighted cross-cutting barriers (time pressure, fragmented handovers, role ambiguity) and pragmatic enablers (brief protected huddles, shared handover templates, clearer task ownership). Overall, variability in interprofessional readiness and psychological safety appeared to be driven more by profession-linked working conditions and organization-wide routines than by ward or unit type. Small-scale, locally adapted quality improvement efforts that protect routine interprofessional touchpoints, standardize handovers, and clarify role ownership warrant prospective evaluation to strengthen collaborative practice in psychiatric care.

## Introduction

Psychiatric care spans acute symptom management to long-term rehabilitation. Interprofessional collaboration among physicians, nursing staff, occupational therapists, psychologists, and psychiatric social workers is therefore central to patient safety, clinical effectiveness, and efficiency (Fleury et al., 2019; Liberman et al., 2001; Reeves et al., 2013; World Health Organization, 2010). In psychiatric hospitals, the quality of collaboration depends not only on individual clinicians’ skills but also on how teams are organized and supported. Conceptual frameworks of interprofessional education highlight shared learning outcomes, role clarification, and reflection as foundations for sustainable collaborative practice (Thistlethwaite, 2012).

Beyond individual dispositions, team climate provides a key psychosocial context for learning and reliable care processes in clinical teams. Team climate includes dimensions such as Participative Safety, Vision, Task Orientation, and Support for Innovation (Anderson & West, 1998; Edmondson, 1999). Team climates that support speaking up, shared goals, and high task standards have been linked to better coordination and safer care. Systematically assessing readiness for interprofessional collaboration and team climate in psychiatric settings, and characterizing how these constructs vary by profession and care setting, is therefore essential for identifying quality-improvement targets and designing context-appropriate interventions.

The Readiness for Interprofessional Learning Scale–Professional version (RIPLS-Pro) assesses clinicians’ attitudinal readiness for interprofessional learning and collaboration across domains such as teamwork, professional identity, and roles and responsibilities (Parsell & Bligh, 1999). The Japanese adaptation of the RIPLS has demonstrated acceptable reliability and cultural validity (Oishi et al., 2017). The Team Climate Inventory (TCI) conceptualizes team functioning across the four dimensions of Vision, Participative Safety, Task Orientation, and Support for Innovation and captures the shared psychosocial context of teams; it has shown reliability and validity in healthcare settings, including those in Japan (Anderson & West, 1998; Onishi, 2013). In this study, we used the RIPLS-Pro total score to index interprofessional readiness at the individual level and focused a priori on the TCI Participative Safety subscale as a key indicator of psychological safety and inclusive voice, which are central to interprofessional collaboration. Descriptive statistics for the other TCI subscales (Vision, Task Orientation, and Support for Innovation) were obtained to characterize the broader team climate in psychiatric care.

Prior work has often focused on learners in interprofessional education contexts (Agreli et al., 2019; Barr, 2015; Parsell & Bligh, 1999; Reeves et al., 2013) or on teams in general hospital settings, which limits transferability to psychiatry (Anderson & West, 1998). Moreover, although profession and placement often covary with age and clinical experience, analyses have seldom adjusted for these confounders. As a result, it remains unclear whether observed differences reflect role-specific factors or career stage. Few studies have jointly examined clinicians’ readiness for interprofessional collaboration and team climate or compared these constructs across both profession and care setting in psychiatric hospitals. Studies are therefore needed that profile interprofessional readiness and team climate among clinicians working in psychiatric hospitals while accounting for basic demographic and experiential differences.

Accordingly, we conducted a single-site cross-sectional survey at a specialist psychiatric hospital in Japan. Using the RIPLS-Pro and the TCI, we examined whether interprofessional readiness and team climate differ across professions (physicians, nursing staff, and allied health professionals) and care settings (outpatient/daycare, acute psychiatric, and chronic care) after adjusting for age and years of clinical experience. We also examined whether profession-by-setting interactions explained additional variance in these outcomes and used brief open-ended comments to contextualize the quantitative patterns. We set two confirmatory aims: (1) to test whether RIPLS-Pro total scores differ by profession after adjustment for age and clinical experience and (2) to test whether TCI Participative Safety subscale scores differ by profession under the same adjustment. Differences in care settings and the profession-by-setting interaction were prespecified as secondary outcomes and interpreted cautiously because of sparse cells. Descriptive analyses of the other TCI subscales (Vision, Task Orientation, and Support for Innovation) were conducted to further characterize the team climate.

## Materials and Methods

### Study Design

We conducted a single-site cross-sectional observational study using a self-administered questionnaire.

### Study Setting

This study was conducted at a single, stand-alone psychiatric hospital in Japan, comprising four primary clinical settings: (1) an outpatient/daycare unit, (2) an acute psychiatric ward, (3) a dementia care ward, and (4) a long-term care ward. The characteristics of each setting, including patient population, mean Global Assessment of Functioning scores, primary clinical roles, number of beds, and median length of stay, are summarized in Supplementary Table S1 (for context only; these variables were not included in the inferential models).

The outpatient/daycare unit provides longitudinal support and social reintegration services for patients with stable chronic psychiatric conditions rather than acute care. In the three months preceding the survey, the mean daily attendance was 32 ± 3.5 patients. This figure is provided for descriptive comparison with the bed counts of inpatient wards because this unit has no inpatient beds.

For inferential analyses, the dementia and long-term care wards were combined a priori and treated as a single chronic care setting. Accordingly, the analyses used three settings: outpatient/daycare, acute psychiatric, and chronic care (combined dementia and long-term care).

### Participants

Eligible participants were all healthcare professionals employed at the hospital in the target professions at the time of the survey (N = 107): physicians (n = 19); nursing staff, comprising registered nursing staff (n = 59) and assistant nursing staff (n = 11); and allied health professionals, comprising occupational therapists (n = 9), certified psychologists (n = 3), and mental health social workers (n = 6). Participation was voluntary and anonymous.

For inferential analyses, the final analytic sample comprised 71 complete cases (overall response rate, 66.4%). By clinical setting, participants were distributed as follows: outpatient/daycare (n = 16), acute psychiatric ward (n = 19), and chronic care (combined dementia and long-term care; n = 36). By profession, the analytic sample included physicians (n = 13), nursing staff (n = 37), and allied health professionals (n = 21). The analytic sample size reflects complete-case availability for the primary outcomes.

### Data Collection

The study was conducted from March to April 2025. An anonymous paper-based self-administered questionnaire was distributed during an on-site research briefing. Participants were instructed to seal the completed forms and deposit them in a locked collection box located in the hospital within the subsequent four weeks. Multiple reminders were posted via the hospital’s internal communication network during the data collection period to maximize participation. No financial or material incentives were offered.

### Measurement Instruments

Two validated instruments were used in this study: the TCI and RIPLS-Pro.

#### RIPLS-Pro

The RIPLS-Pro was used to assess practicing clinicians’ readiness for interprofessional collaboration. It comprises four subscales: IPL in Practice, Patient-Centeredness, IPL in Education, and Sense of Interprofessional Identity. Each item is rated on a 5-point Likert scale (1 = strongly disagree to 5 = strongly agree). Subscale and total scores were computed as the sum of the item scores, with higher values indicating greater IPL readiness. Several items were reverse-scored according to the validated Japanese version. The reliability and validity of the Japanese RIPLS-Pro have been confirmed in a previous study (Oishi et al., 2017).

Regarding the subscales, IPL in Practice reflects attitudes toward interprofessional collaboration in clinical settings. Patient-Centeredness captures the perceived importance of patient-centered care. IPL in Education represents one’s views on the necessity of interprofessional education for qualified professionals. Finally, Sense of Interprofessional Identity indicates awareness of one’s professional role and identity in relation to other professions.

#### Team Climate Inventory

The TCI was used to evaluate the psychosocial climate of clinical teams across four subscales: Vision, Participative Safety, Support for Innovation, and Task Orientation. The instrument comprises 38 items rated on a 5-point Likert scale (1 = strongly disagree to 5 = strongly agree). Subscale and total scores were calculated as the sum of item scores, with higher values indicating a more favorable team climate. Reverse-coded items were scored according to the official manual where applicable. The reliability and validity of the Japanese version of the TCI have been confirmed in a previous study (Onishi, 2013).

Regarding the subscales, Vision represents shared objectives and aspirations that provide meaning and direction to teamwork. Participative Safety reflects a psychologically safe environment in which team members can express opinions and participate in decision-making without interpersonal risk. Support for Innovation captures the extent to which new ideas and change initiatives are encouraged and supported. Finally, Task Orientation denotes members’ commitment to high-quality performance through evaluation and continuous improvement aligned with team goals.

#### Open-ended Items

Participants responded to open-ended questions regarding the challenges they face related to intra- and interprofessional collaboration. These responses were collected to contextualize the quantitative findings (see the Data Analysis section).

### Data Analysis

Internal consistency was assessed using Cronbach’s α for the RIPLS-Pro total and subscales and for each TCI subscale; coefficients are reported in the Supplementary Materials. Descriptive statistics for continuous variables are presented as mean ± standard deviation (SD). Baseline differences in age and years of clinical experience across professions and care settings were examined using one-way analysis of variance (ANOVA). Pearson’s χ² tests were used to compare differences in gender distributions.

The primary outcome was the RIPLS-Pro total score. The secondary outcome was the TCI Participative Safety subscale score. Descriptive analyses of the other TCI subscales (Vision, Task Orientation, and Support for Innovation) were conducted to further characterize the team climate, and their results are provided in the Supplementary Materials.

Primary inferential analyses used analysis of covariance (ANCOVA) models with profession (physicians, nursing staff, allied health professionals) and care setting (outpatient/daycare, acute psychiatric ward, chronic care) as fixed factors and age and years of clinical experience as continuous covariates. The main analyses focused on the main effects of profession and care setting on each outcome. The profession × setting interaction was prespecified as of secondary interest because of sparse cells in several combinations and is therefore reported only in the Supplementary Materials.

Estimated marginal means (EMMs) with 95% confidence intervals (CIs) for each profession and care setting were reported and evaluated at the sample means of the covariates. When a main effect was statistically significant, Bonferroni-adjusted pairwise EMM contrasts were conducted. Effect sizes were summarized using partial η^²^. Model assumptions were examined using residual diagnostics. The homogeneity of variances was assessed using Levene’s test, and the homogeneity of regression slopes was evaluated by adding covariate × factor interaction terms to the models. All analyses were based on the same complete-case sample because no missing values remained in the outcomes or covariates after data cleaning. All tests were two-sided with α = 0.05. Analyses were performed using IBM SPSS Statistics version 21 (IBM Corp., Armonk, NY, USA).

Sensitivity analyses were conducted using ANCOVA models that included the profession × setting interaction term in the main specification, mirroring the primary analytic approach. The results are provided in the Supplementary Materials.

### Qualitative Analysis of Open-Ended Responses

The participants answered two open-ended questions regarding what helped and hindered intra- and interprofessional collaboration in their daily work. We assessed their responses using directed content analysis grounded in the RIPLS-Pro and TCI subscales. Two researchers independently coded all responses. Comments were brief, and coding was used descriptively to contextualize the quantitative findings; therefore, formal interrater reliability was not calculated. Any discrepancies were resolved through consensus. Themes were summarized by profession and care setting, and representative quotations illustrating each theme are presented in the Results section.

### Ethical Considerations

This study was approved by the Ethics Committee for Research Involving Human Subjects at Nihon Fukushi University (approval number: 24-038-02). All eligible staff members received written information about the study. Participation was voluntary and anonymous, and written informed consent was obtained before the survey. Participants could withdraw at any time without disadvantages. No financial or material incentives were provided. Questionnaires were returned in sealed envelopes to a locked collection box.

All data were de-identified prior to analysis and stored on password-protected institutional servers that were accessible only to the research team. Participants also provided consent for the use of anonymized, de-identified excerpts from their free-text comments in publications. The study adhered to the principles of the Declaration of Helsinki and relevant institutional and national guidelines. Reporting follows the Strengthening the Reporting of Observational Studies in Epidemiology (STROBE) guidelines to ensure transparency and rigor.

## Results

### Participants and Settings

We analyzed data from 71 healthcare professionals (physicians, n = 13; nursing staff, n = 37; allied health professionals, n = 21). Participants worked in outpatient/daycare services (n = 16), an acute psychiatric ward (n = 19), or chronic care wards (combined dementia and long-term care; n = 36). In this study, “allied health” refers to mental health social workers, occupational therapists, and certified psychologists. All outcomes and covariates were complete; thus, the same sample (N = 71) was used for all analyses.

Table 1 presents the counts by care setting and profession, including the setting × profession cross-tabulation. Contextual characteristics of service settings are summarized in Supplementary Table S1. Age and years of clinical experience were entered as covariates in all adjusted analyses, and EMMs were evaluated at their sample means. Assumption checks indicated no material violations of residual normality, homogeneity of variances (Levene), or homogeneity of regression slopes.

**Table 1.** Sample characteristics and cross-tabulation by care setting and profession (N = 71)

| Care setting | Physicians | nursing staff | Allied health | Total |
| --- | --- | --- | --- | --- |
| Outpatient/daycare | 2 | 9 | 5 | 16 |
| Acute | 5 | 10 | 4 | 19 |
| Chronic care | 6 | 18 | 12 | 36 |
| Total | 13 | 37 | 21 | 71 |

Key demographics (overall)
- Age (years), mean ± SD: 45.1 ± 11.8 - Years of clinical experience, mean ± SD: 10.0 ± 11.0 - Female, n (%): 38 (53.5)
Notes. Only variables used in the models are shown. Age and years of clinical experience were entered as covariates. Estimated marginal means (EMMs) were evaluated at their sample means. DT, dementia treatment; LTC, long-term care.

### Internal Consistency of Measures

Cronbach’s α indicated acceptable reliability for both instruments (RIPLS-Pro total α = .79; TCI total α = .80). The scale- and subscale-level contributions, including those of the TCI Participative Safety subscale, are reported in Supplementary Table S2.

### Primary Outcome: RIPLS-Pro Total Score

In the ANCOVA model, age (F = 14.33, p < .001, partial η^²^ = .220) and years of clinical experience (F = 13.95, p < .001, partial η^²^ = .215) were independently associated with RIPLS-Pro total scores. Profession had a significant main effect (F = 4.36, p = .018, partial η^²^ = .140), whereas the main effect of care setting (F = 2.27, p = .113, partial η^²^ = .075) was not significant. Model fit was R² = .336 (adjusted R² = .226). The profession-wise adjusted means are shown in Figure 1, and full ANCOVA results are provided in Supplementary Table S3. EMMs [95% CIs] were as follows: physicians 147.96 [138.75–157.18], nursing staff 139.83 [134.30–145.35], and allied health professionals 152.34 [145.60–159.09].

**Figure 1.**
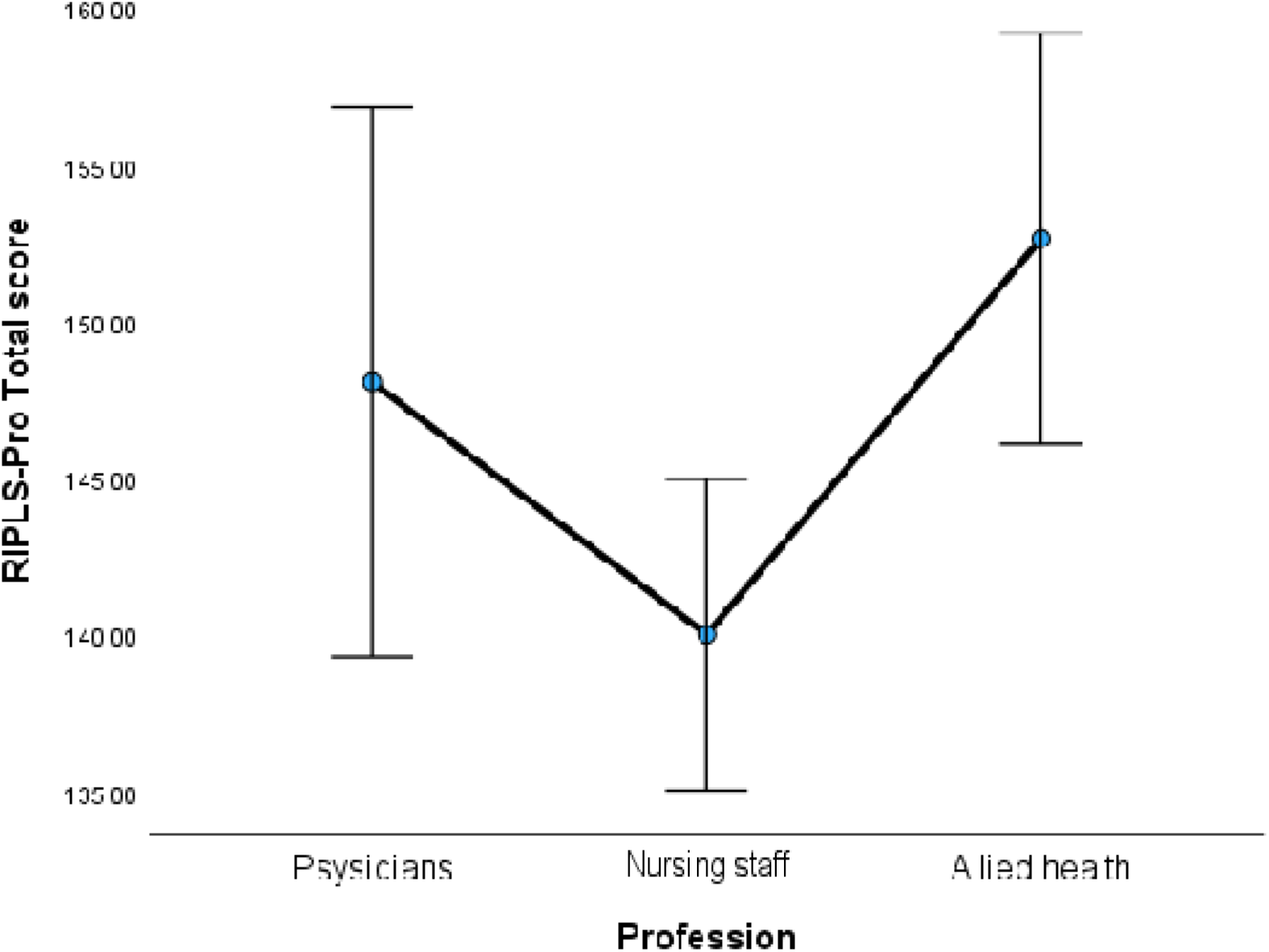
Estimated marginal means (95% CI) — RIPLS-Pro Total by profession. Values used (EMM, 95% CI): Physician 147.96 [138.75–157.18]; Nursing staff 139.83 [134.30–145.35]; Allied health 152.34 [145.60–159.09].

Bonferroni-adjusted pairwise EMM contrasts showed that allied health professionals > nursing staff; physicians vs. allied health professionals and physicians vs. nursing staff were not significant (see Supplementary Table S4).

### Secondary Outcome: TCI Participative Safety

Age was significantly associated with the Participative Safety subscale (F = 5.92, p = .017, partial η^²^ = .095); however, the association with years of experience was not significant (F = 3.53, p = .065, partial η^²^ = .058). Profession showed a significant main effect (F = 6.10, p = .005, partial η^²^ = .175), whereas the main effect of care setting (F = 0.31, p = .734, partial η^²^ = .011) was not significant. Model fit was R² = .385 (adjusted R² = .285). The profession-wise adjusted means are shown in Figure 2, and full ANCOVA results are provided in Supplementary Table S5. EMMs [95% CIs] were consistent with the pattern observed for RIPLS-Pro as follows: physicians 47.04 [43.63–50.46], nursing staff 42.52 [40.47–44.57], and allied health professionals 47.54 [45.03–50.04]. Bonferroni-adjusted pairwise EMM contrasts indicated that allied health professionals > nursing staff. The remaining pairs were not significant (see Supplementary Table S4). Sensitivity analyses including the profession × setting interaction showed no significant interaction effects for either the RIPLS-Pro total or TCI Participative Safety subscale scores (see Supplementary Tables S3 and S5).

**Figure 2.**
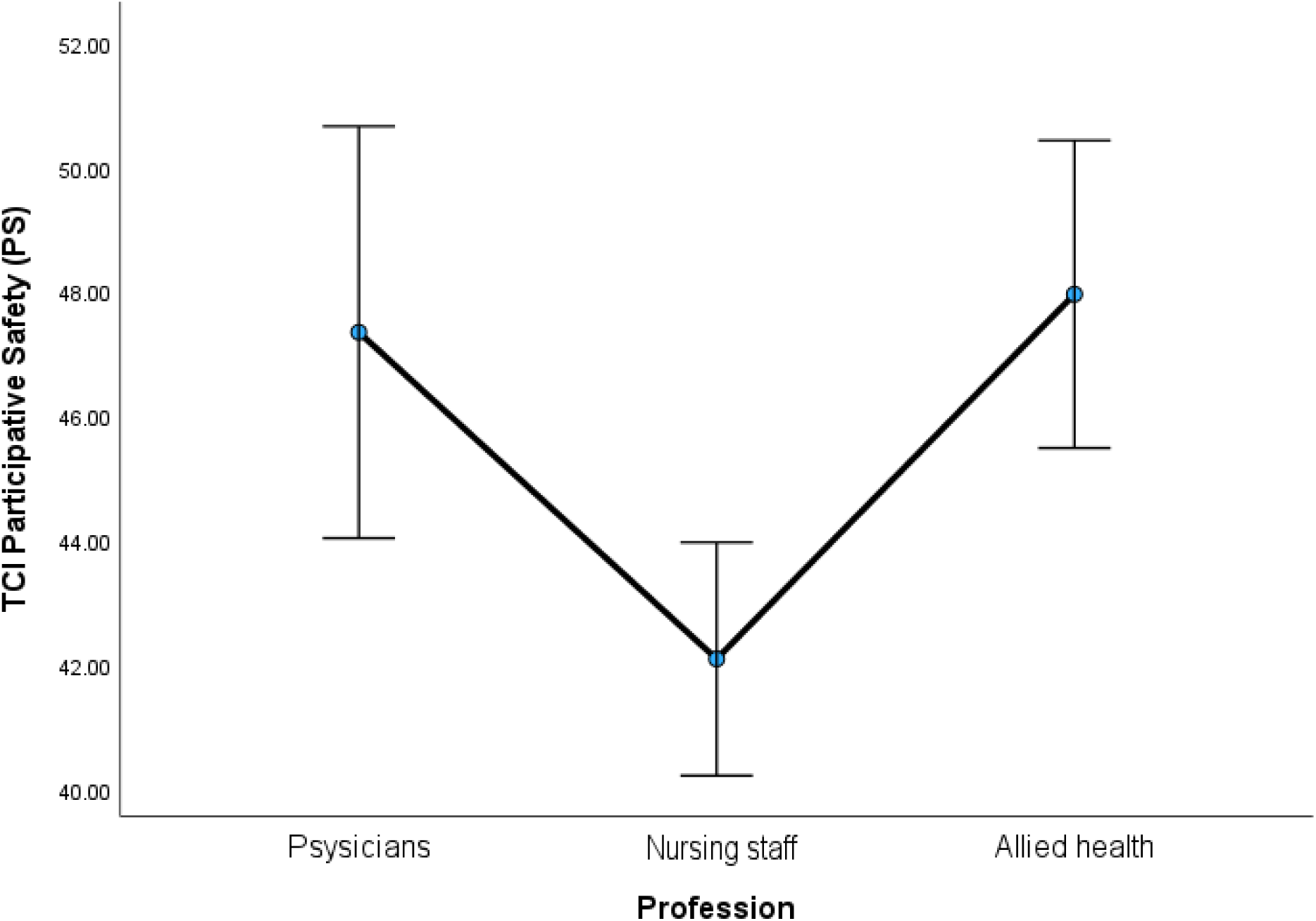
Estimated marginal means (95% CI) — TCI Participative Safety (PS) by profession. Values used (EMM, 95% CI): Physician 47.04 [43.63–50.46]; Nursing staff 42.52 [40.47–44.57]; Allied health 47.54 [45.03–50.04].

### Qualitative Findings

A directed content analysis of free-text comments was conducted to organize the material into three categories aligned conceptually with the RIPLS-Pro and TCI constructs: Enablers, Barriers, and Actions and Support. Representative quotations are provided below; additional examples are available upon request.

#### Enablers

Across professions, respondents emphasized routine touchpoints and simple, shared formats for information exchange (brief huddles/mini-conferences, standardized handover notes, and agreed messaging channels) as facilitators of teamwork. One member of the nursing staff stated, “The morning mini-conference anchors a shared understanding for the day.”

#### Barriers

The most common obstacles were related to time pressure and information flow. Physicians highlighted scheduling constraints that reduced overlap for team discussion, while allied health staff underscored ambiguity over task ownership during care transitions, mapping to role clarity and psychological safety for speaking up. According to one physician in the outpatient/daycare unit, “Clinic schedules leave little overlap for team discussion.”

#### Actions and Support

Proposed solutions centered on clarifying ownership (e.g., designating a lead for family inclusive care conferences), creating short, predictable huddles, and adopting lean sharing templates. Educational supports included interprofessional case reviews and onboarding-level interprofessional education. An allied health professional who worked in chronic care stated, “A simple protocol that assigns a lead and a single point of contact would help.”

## Discussion

This cross-sectional study yielded two main findings. First, after adjusting for age and years in practice, both the RIPLS-Pro total score and the TCI Participative Safety subscale score differed systematically by profession: allied health professionals scored the highest, followed by physicians and then nursing staff. Second, clinical setting (outpatient/daycare, acute, and long-term care) and the profession-by-setting interaction did not explain additional variance. Taken together, these results suggest that interprofessional readiness and psychological safety in psychiatric care are shaped more by profession-linked working conditions and organization-wide routines than by ward or unit type.

Regarding RIPLS-Pro scores, adjusted means showed a clear gradient (allied health professionals 152.34 [95% CI 145.60–159.09], physicians 147.96 [138.75–157.18], nursing staff 139.83 [134.30–145.35]) with moderate model fit (R² = .336; adjusted R² = .226). Age was positively associated with RIPLS-Pro scores, consistent with evidence from previous studies indicating that IPL readiness tends to increase with cumulative clinical experience and repeated exposure to collaborative practice (Johnson et al., 2017). Differential exposure to structured interprofessional activities provides a plausible explanation for this profession gradient. In many psychiatric hospitals, allied health professionals regularly participate in scheduled case conferences and goal-setting meetings, whereas nursing staff often shoulder broad task loads and intensive intra-professional coordination under time pressure (Liberman et al., 2001). These organizational conditions, including how meetings are scheduled, how handovers are structured, and who is responsible for updating shared plans, are likely to shape readiness for collaboration more strongly than inherent professional traits.

Regarding TCI Participative Safety scores, the adjusted means again favored allied health professionals (47.54 [45.03–50.04]) and physicians (47.04 [43.63–50.46]) over nursing staff (42.52 [40.47–44.57]), with similar model fit (R² = .385; adjusted R² = .285). Participative safety reflects psychological safety and inclusive communication within teams (Edmondson, 1999). Previous studies have indicated that such climate factors depend more on leadership style, trust, and shared norms than on physical work settings (Anderson & West, 1998; Onishi, 2013). The absence of significant setting effects in this study supports this view. Differences in profession-linked routines, expectations, and leadership dynamics appear to outweigh environmental differences among outpatient/daycare, acute, and long-term care units.

Qualitative responses help interpret these patterns. Participants most frequently cited time pressure, fragmented handovers, and role ambiguity as barriers. Prior studies on interprofessional collaboration in psychiatric and general healthcare have also documented these issues (Kiger et al., 2021; Reeves et al., 2013). The suggested facilitators, including brief protected touchpoints, standardized handover templates, and clearer ownership of cross-disciplinary tasks, align with evidence that structured communication and defined roles enhance team coordination and safety (Agreli et al., 2019; Fleury et al., 2019). The recurrence of these themes across all clinical settings reinforces the interpretation that systemic, organizational factors, rather than ward-specific cultures, underlie much of the observed variability in readiness and team climate. These findings are consistent with work in local mental health service networks showing that leadership, role clarity, and organizational factors are key correlates of interprofessional collaboration (Ndibu Muntu Keba Kebe et al., 2019). Together, these comments suggest that cross-cutting organizational routines and constraints, rather than ward type alone, are central leverage points for strengthening interprofessional readiness and participative safety.

### Implications for Interprofessional Practice and Education

This study’s findings have several implications for interprofessional development in psychiatric care. Lower scores among nursing staff suggest a need to support their role as coordinators and information hubs within interprofessional psychiatric teams, for example through targeted training and protected time for participation in case discussions. Higher scores among allied health professionals may reflect routines and collaborative practices that could be made more visible and shared across professions in interprofessional education and in-service training. The identified cross-cutting barriers and facilitators, including time allocation for brief huddles, standardized handovers, and clearer role ownership, provide concrete, low-cost targets for locally adapted quality-improvement efforts that could strengthen collaborative practice without major structural reforms.

### Limitations

This study had several limitations. It was a single-site, cross-sectional study with a modest sample size (N = 71), which limits generalizability and precludes causal inference. All outcomes were self-reported, raising the possibility of common method variance and social desirability bias. Behavioral indicators and patient-level outcomes were not assessed. Furthermore, residual confounding may persist, considering that data on workload, staffing ratios, supervisory structures, and local leadership styles were unavailable. Future research should use multi-site longitudinal designs incorporating fidelity and outcome measures to evaluate whether small, locally adapted adjustments in scheduling, handover structures, role clarity, and facilitation can sustainably improve interprofessional readiness, team climate, and ultimately, patient and family outcomes in psychiatric care (Agreli et al., 2019; Reeves et al., 2013).

## Conclusion

In this single-site psychiatric hospital, interprofessional readiness (RIPLS-Pro) and team climate (TCI Participative Safety) differed by profession but not by care setting after adjusting for age and experience. Allied health professionals scored the highest, followed by physicians and nursing staff. Free-text comments highlighted cross-cutting barriers (time pressure and role ambiguity) and feasible enablers (brief huddles and standardized handovers). These findings suggest that collaborative quality is shaped more by profession-linked routines and organization-wide practices than by unit type. Future prospective multisite studies should examine whether locally tailored changes to scheduling, handovers, and role clarity can improve team climate and care outcomes.

## Supporting information

Supplementary Table S1. Characteristics of surveyed psychiatric hospital work settings

Supplementary Table S3. ANCOVA results for RIPLS-Pro total scores (covariates: age, years of experience)

Supplementary Table S4. Pairwise EMM contrasts by profession (Bonferroni-adjusted)

Supplementary Table S5. ANCOVA results for TCI Participative Safety subscale scores (covariates: age, years of experience)

## Notes on contributors

**Yasuhisa Nakamura** is a Lecturer in the Course of Occupational Therapy, Department of Rehabilitation, Faculty of Health Sciences, Nihon Fukushi University. His research interests include psychiatric rehabilitation and interprofessional education in mental health.

**Mayumi Yoshikawa** is an Assistant Professor in the Faculty of Social Welfare, Nihon Fukushi University. Her research interests are in mental health social work and psychosocial support for individuals with mental health disorders.

**Koshi Terashita** is a Registered Nurse and Director of Nursing at Minamichita Hospital, a psychiatric hospital in Japan. Terashita’s clinical interests focus on psychiatric nursing and the delivery of high-quality care in inpatient mental health settings.

**Miki Fukushima** is a Lecturer in the Department of Nursing, College of Life and Health Sciences, Chubu University. Her research interests include psychiatric nursing, ethical dilemmas in mental health care, and nursing education.

**Chizuru Tsubonouchi, PhD,** is an Assistant Professor in the Department of Nursing, Faculty of Nursing, Nihon Fukushi University. Her research interest includes narrative approaches to psychiatric nursing (e.g., the patient-authored medical records) and mental health care for the patients and their families.

## Funding

This research received no specific grant from any funding agency in the public, commercial, or not-for-profit sectors.

## Conflict of Interest Disclosure

The authors report there are no conflicts of interest to declare.

## Data Availability Statement

The data that support the findings of this study are available from the corresponding author upon reasonable request.

## Ethical Approval Statement

The study protocol was approved by the Clinical Research Ethics Committee of Nihon Fukushi University, Japan (approval no: 24-038-02).

## Participant Consent Statement

All participants were fully informed about the nature and purpose of the study, and written informed consent was obtained prior to participation.

## CLINICAL TRIAL REGISTRATION

The trial was registered with the University Hospital Medical Information Network (UMIN) Clinical Trials Registry (UMIN000057989).

## Declaration of Generative AI and AI-Assisted Technologies in the Writing Process

During the preparation of this manuscript, the authors used ChatGPT (OpenAI, large language model; accessed in 2025) to assist with improving the clarity and readability of the English language. ChatGPT was not used to generate, analyze, or interpret any data. All final editing and content decisions were made by the authors, who take full responsibility for the content of the manuscript.

## Notes

### Competing Interest Statement

The authors have declared no competing interest.

### Clinical Trial

UMIN000057989

### Funding Statement

This study did not receive any funding

