## Supplementary Table S1. Characteristics of surveyed psychiatric hospital work settings for "Interprofessional Readiness and Team Climate in Psychiatric Care: Profession-Specific Differences in a Japanese Psychiatric Hospital"

| **Work setting** | **Principal diagnoses** | **Mean GAF (±SD)** | **Primary Role** | **Beds** | **Median Length of Stay (range)** |
| --- | --- | --- | --- | --- | --- |
| Outpatient/daycare unit | Schizophrenia | 45 ± 9.1 | Outpatient treatment, social reintegration support | ― | ― |
| Acute psychiatric ward | Mood disorder,  Schizophrenia | 32 ± 8.0 | Short-term treatment for acute symptoms | 44 | 55 (40–60) |
| chronic care ward | Schizophrenia, Dementia | 30 ± 10 | Long-term care for chronic cases | 120 | 687 (241–3714) |
| Dementia care ward | Dementia | 28 ± 7.2 | Specialized treatment for dementia | 54 | 319 (232–2754) |

**Notes.** GAF, Global Assessment of Functioning. Dashes indicate “not applicable” or “not routinely recorded.” For modeling, chronic care wards were combined into a single “chronic care” category. These contextual variables were not included as covariates, but are provided to describe service settings.
