## Supplemental Data 1 for "Interprofessional Readiness and Team Climate in Psychiatric Care: Profession-Specific Differences in a Japanese Psychiatric Hospital"

**Supplementary Table S2. Reliability summary (Cronbach’s alpha) and subscale contributions**

**A. RIPLS-Pro**

| **Scale / Subscale** | **Cronbach’s α (total scale)** | **Corrected subscale–total correlation** | **α if subscale deleted** |
| --- | --- | --- | --- |
| RIPLS-Pro Total | .79 | — | — |
| IPL in Practice | — | .934 | .605 |
| Patient-centeredness | — | .574 | .804 |
| IPL in Education | — | .809 | .747 |
| Sense of Interprofessional Identity | — | .506 | .791 |

*Source for α (overall): SPSS reliability output (α = 0.788). CITC/α-if-deleted from the same run.*

**B. Team Climate Inventory (TCI)**

| Scale / Subscale | Cronbach’s α (total scale) | Corrected subscale–total correlation | α if subscale deleted |
| --- | --- | --- | --- |
| TCI (4 Subscale aggregated) | .80 | — | — |
| Vision | — | .396 | .854 |
| Participative Safety（PS） | — | .685 | .714 |
| Support for Innovation | — | .733 | .705 |
| Task Orientation | — | .697 | .710 |

*Source for α (overall across Subscale): SPSS reliability output (α = 0.800). Subscale-level CITC/α-if-deleted from the same run.*
