## Supplementary Table S3. ANCOVA results for RIPLS-Pro total scores (covariates: age, years of experience) for "Interprofessional Readiness and Team Climate in Psychiatric Care: Profession-Specific Differences in a Japanese Psychiatric Hospital"

| Effect | df | F | p | Partial η² |
| --- | --- | --- | --- | --- |
| Age | 1 | 14.33 | <.001 | .220 |
| Years of experience | 1 | 13.95 | <.001 | .215 |
| Care setting (3 levels) | 2 | 2.27 | .113 | .075 |
| Profession (3 levels) | 2 | 4.36 | .018 | .140 |
| Setting × Profession | 4 | 1.76 | .149 | .115 |

Model fit: R² = .336 (adjusted R² = .226). Type-III sums of squares.
