## Supplementary Table S4. Pairwise EMM contrasts by profession (Bonferroni-adjusted) for "Interprofessional Readiness and Team Climate in Psychiatric Care: Profession-Specific Differences in a Japanese Psychiatric Hospital"

| Outcome | Contrast (A−B) | EMM difference | SE | 95% CI | df | Adjusted *p* |
| --- | --- | --- | --- | --- | --- | --- |
| RIPLS-Pro total | Allied health – Nursing staff | +12.517 | 4.493 | [1.451, 23.583] | 60 | .021 |
| Physician − allied health | −4.379 | 5.966 | [−19.072, 10.315] | 60 | 1.000 |  |
| Physician − Nursing staff | 8.138 | 5.226 | [−4.732, 21.009] | 60 | .374 |  |
| TCI—Participative Safety (PS) | Allied health – Nursing staff | +5.022 | 1.666 | [0.918, 9.125] | 60 | .011 |
| Physician − allied health | −0.494 | 2.212 | [−5.943, 4.955] | 60 | 1.000 |  |
| Physician − Nursing staff | 4.527 | 1.938 | [−0.245, 9.300] | 60 | .069 |  |

Notes: Positive differences favor the first group (A). EMMs are evaluated at the sample means of age and years of clinical experience (45.1 and 10.0, respectively). Models are ANCOVA with age and years of experience as covariates; Type-III SS; Bonferroni adjustment for multiple pairwise tests.
