## Supplementary Table S5. ANCOVA results for TCI Participative Safety subscale scores (covariates: age, years of experience) for "Interprofessional Readiness and Team Climate in Psychiatric Care: Profession-Specific Differences in a Japanese Psychiatric Hospital"

| Effect | df | F | p | Partial η² |
| --- | --- | --- | --- | --- |
| Age | 1 | 5.92 | .017 | .095 |
| Years of experience | 1 | 3.53 | .065 | .058 |
| Care setting (3 levels) | 2 | 0.31 | .734 | .011 |
| Profession (3 levels) | 2 | 6.10 | .005 | .175 |
| Setting × Profession | 4 | 1.20 | .324 | .079 |

*Model fit:* R² = .385 (adjusted R² = .285). Type-III sums of squares.
